# Association Between Life’s Essential 8 and Atherogenic Index of Plasma in Adults: Insights from NHANES 2007-2018

**DOI:** 10.1101/2024.09.16.24313778

**Authors:** Long-hui Xu, Kai-wen Ding, Guo-dong Yang, Xiao-xuan Han, Xiao Cong, Rong-hui Wang, Xin-ru Liu, Na Li, Cui-ping Xu

**Author notes:** Corresponding Author: Cui-ping Xu, PhD, Chief Nurse. These authors contributed equally to this work and should be considered co-first authors (Long-hui Xu, Kai-wen Ding). **Author Note:** Longhui Xu, MSN in reading, Student. Kai-wen Ding, MSN, Supervisor nurse. Guo-dong Yang, MSN in reading, Student. Xiao-xuan Han, MSN in reading, Student. Xiao Cong, MSN in reading, Student. Rong-hui Wang, MSN in reading, Student. Xin-ru Liu, MSN in reading, Student. Na Li, MSN in reading, Student.

## Abstract

**Objectives:** This study aimed to investigate the relationship between Life’s Essential 8 (LE8) and the Atherogenic Index of Plasma (AIP).

**Methods:** We conducted an analysis of data from 8,215 U.S. adults aged 20 years and older, utilizing the National Health and Nutrition Examination Survey data from 2007 to 2018. Based on LE8 scores, Cardiovascular Health (CVH) was stratified into three levels—low, moderate, and high—while AIP was categorized into four risk levels: extremely low (AIP<-0.3), low (−0.3≤AIP<0.1), medium (0.1≤AIP<0.24), and high (AIP≥0.24). Weighted ordinal logistic regression analysis was utilized to examine the association between CVH scores and AIP risk levels, adjusting for potential confounding variables.

**Results:** A significant inverse correlation exists between CVH scores and AIP risk levels (*OR*=0.51, *95%CI*: 0.49-0.54, *P*<0.001). Higher CVH scores were associated with lower AIP risk levels, while lower CVH scores corresponded to elevated AIP risk levels. Notably, improvements in specific CVH components—such as Body Mass Index and Blood Lipids—exhibited a strong relationship with reductions in AIP risk levels.

**Conclusions:** Enhancing CVH is vital for effectively reducing AIP risk levels, thus underscoring the critical importance of health management strategies in the prevention of cardiovascular diseases.

## 1. Introduction

Cardiovascular Disease (CVD), primarily encompassing ischemic heart disease and stroke, has emerged as an increasingly severe public health challenge on a global scale.^1^ According to statistical data, the number of deaths attributable to CVD worldwide rose from 12.4 million in 1990 to 19.8 million in 2022, with Eastern Europe exhibiting the highest age-standardized mortality rate, reaching 553 deaths per 100,000 individuals.^2,3^ In light of this alarming trend, the American Heart Association (AHA) formally introduced the concept of Cardiovascular Health (CVH) in 2010, aimed at evaluating and enhancing cardiovascular well-being among individuals and populations by quantifying specific health metrics. This initiative seeks to mitigate the risk of CVD, improve quality of life, extend longevity, reduce healthcare expenditures, and foster public health.^4,5^ The initial framework of CVH, referred to as “Life’s Simple 7” (LS7), comprises seven principal cardiovascular risk factors: diet, physical activity, current smoking, body mass index (BMI), total cholesterol, blood pressure, and fasting plasma glucose. Each metric is categorized into three levels—poor, intermediate, or ideal—based on clinical cut points, with an overall CVH status quantified by a score ranging from 0 to 14. ^4,6^

As research has progressed, the LS7 framework has been found to exhibit several limitations in the assessment of CVH, including insufficient sensitivity to individual heterogeneity, an incomplete representation of healthy dietary characteristics within the dietary metric, a lack of comprehensive consideration of health behaviors, and potential misclassification biases.^7,8^ To address these shortcomings, the AHA has recently undertaken a comprehensive expansion of the CVH definition, introducing the “Life’s Essential 8” (LE8) framework. This updated model incorporates a new metric for sleep health and refines the quantification of the original indicators, adopting a continuous scoring system ranging from 0 to 100 to more accurately evaluate an individual’s cardiovascular well-being.^7,9^ Subsequently, the work of López-Bueno et al.^10^ has estimated the global prevalence of the eight key factors comprising the LE8, further corroborating the efficacy and utility of the LE8 framework in CVH assessment and laying the foundation for further exploration of the relationships between the LE8 components and CVH indicators.

The Atherogenic Index of Plasma (AIP) is a novel biomarker that quantifies an individual’s risk of atherosclerosis. It is calculated as the logarithm of the ratio of triglycerides (TG) to high-density lipoprotein cholesterol (HDL-C), with the aim of effectively predicting the occurrence and prognosis of CVD, as well as guiding clinical interventions.^11–13^ Previous research has demonstrated that AIP is not only associated with the risk of CVD^14^, but also correlates with various LE8 metrics, including BMI^15^, Healthy Eating Index-2015 (HEI-2015)^16^, physical activity^17^, blood lipids^18^, blood glucose^19^, and blood pressure^20^.

Despite the wealth of independent research on LE8 and AIP, the relationship between the two remains inadequately explored, particularly in terms of developing effective assessment models to elucidate the specific impacts of each LE8 component on AIP. Therefore, this study conducted a comprehensive analysis utilizing data from the National Health and Nutrition Examination Survey (NHANES) in the United States, aiming to provide a more precise basis for CVD risk assessment, identify key lifestyle intervention targets, and ultimately enhance the decision-making capabilities of healthcare professionals in health management.

## 2. Methods

### 2.1. Data source and participants selection

The data were sourced from the NHANES database (https://wwwn.cdc.gov/nchs/nhanes/Default.aspx). NHANES, initiated by the Centers for Disease Control and Prevention (CDC), is a comprehensive survey utilizing questionnaires, laboratory tests, and physical examinations. The survey employs a multi-stage, stratified, and clustered probability sampling design to assess the health and nutritional status of a nationally representative sample of the U.S. civilian population, and to determine the prevalence and risk factors of major diseases. All study protocols have been approved by the National Center for Health Statistics Research Ethics Review Board, and written informed consent was obtained from all participants.^21,22^

This study initially included 59,842 participants from six NHANES cycles (2007-2018). After applying exclusion criteria, a total of 8,215 participants were included in the final analysis. Exclusion criteria comprised: (1)insufficient data for calculating the AIP; (2) inadequate data for computing the LE8 score; (3) age below 20 years; (4) incomplete Patient Health Questionnaire-9 (PHQ-9) responses; (5) missing alcohol consumption data; (6) absent Poverty Income Ratio information; (7) lacking education level data; and (8) unknown marital status. (See **Supplementary Figure 1**).

### 2.2. Assessment of CVH by LE8

This study utilized the LE8 framework proposed by the AHA to assess CVH. The LE8 system integrated four health behavior metrics—diet, physical activity, nicotine exposure, and sleep—and four health factor metrics—BMI, blood lipids, blood glucose, and blood pressure. Each metric was evaluated on a 0-100 point scale. The overall CVH score was calculated as the unweighted average of these eight metrics and was categorized into high (80-100), moderate (50-79), and low (0-49) levels in accordance with AHA guidelines.^7^

Diet quality was evaluated using the HEI-2015, which was calculated based on two 24-hour dietary recall surveys from participants and the USDA’s food pattern equivalents database. The HEI-2015 was a comprehensive tool comprising 13 components to assess dietary quality, with a maximum score of 100, where higher scores indicated healthier diets. Nine of these components focused on dietary adequacy (total fruits, whole fruits, total vegetables and legumes, whole grains, dairy, total protein foods, seafood and plant proteins, and fatty acids), while the remaining four assessed dietary moderation (intake of refined grains, sodium, added sugars, and saturated fats). Furthermore, to ensure the reliability and representativeness of blood pressure data, the study employed a standardized method of averaging three measurements. Detailed scoring criteria could be found in **Supplementary Table 1** and **Supplementary Table 2**.

### 2.3 Atherogenic index of plasma

The AIP was mathematically derived from the logarithm of the ratio of triglyceride to high-density lipoprotein cholesterol, both expressed in mmol/L, as follows: AIP=log10[TG(mmol/L) /HDL-C(mmol/L)].^11^ Based on AIP values, participants were stratified into four risk categories: Extremely Low Risk Level (AIP <-0.3), Low Risk Level (−0.3≤ AIP < 0.1), Medium Risk Level (0.1≤ AIP < 0.24), and High Risk Level (AIP ≥0.24).^23^

### 2.4 Covariates

Covariates included age, sex (male and female), race (Mexican American, non-Hispanic White, non-Hispanic Black, and others), Poverty-to-income ratio [categorized as low income (<1.3), medium income (1.3-3.5), and high income (≥3.5)], marital status (divorced/separated/widowed, married/living with partner, and never married), educational level (high school or less, some college, and college graduate or above), presence of depression, and average daily alcohol consumption in the past 12 months.

### 2.5 Statistical analysis

NHANES employed design weights to ensure the representativeness of its data. Since this study utilized data from two 24-hour dietary recalls, the specific weight “Dietary Two-Day Sample Weight (WTDR2D)” was applied according to NHANES guidelines. The WTDR2D weight was constructed based on the two-year cycle sample weight from the MEC, and was further adjusted for (a) additional non-response and (b) distributional differences by day of the week in the collection of dietary intake data. Continuous variables were represented as mean ± standard deviation, while categorical variables were presented as counts (percentages). Group differences were assessed using a weighted linear regression model and a weighted chi-square test.

After adjusting for potential confounding factors, including age, sex, race, education level, marital status, poverty-to-income ratio, 12-month average daily alcohol consumption, and depression, a weighted ordinal logistic regression was employed to examine the association between the CVH score and the AIP risk level. The results were reported as adjusted odds ratios (ORs) with 95% confidence intervals (CIs). To test the proportional odds assumption—where a variable’s coefficients in the model’s different threshold equations are approximately the same or very similar, indicating that it satisfies the proportional odds assumption—a parallelism test was conducted. However, due to the large sample size, the parallelism test was overly sensitive, and even minor differences in coefficients could be statistically identified as violating the assumption. Therefore, this study employed the Wald test of the parallel lines assumption, and sequentially calculating the coefficients of each factor in the model, finding that the three models could be generally considered parallel.

All statistical analyses were performed using R software (version 4.3.2, The R Foundation; http://www.R-project.org) and STATA software (version 18, StataCorp; http://www.stata.com). A significance level of *P*<0.05 was considered statistically significant.

## 3. Result

### 3.1. General characteristics of the study population

**Table 1** presents the general characteristics of the study population based on different AIP risk levels derived from the NHANES 2007-2018 data. Highly significant differences (*P*<0.0001) were observed among participants across the different AIP risk levels in terms of age, sex, race, education level, marital status, poverty-to-income ratio, 12-month average daily alcohol consumption, depression, LE8 score, CVH score, health behaviors score, and health factors score. Compared to the lower risk level group, the high AIP risk level group was characterized by older age, higher male proportion, greater non-Hispanic White representation, predominantly married/living with a partner, lower educational attainment, and reduced 12-month average daily alcohol consumption. Moreover, a notable gradient effect was observed between LE8 scores and AIP risk levels: LE8 scores decreased consistently as AIP risk increased. This trend was most pronounced in the Health Factors dimension, with scores declining from 82.52 in the extremely low AIP risk level group to 56.25 in the high risk AIP level group. However, certain indicators (physical activity, sleep health, and BMI) showed slight improvements in the high AIP risk level group, differing from the overall gradient trend.

**Table 1.** Weighted baseline characteristics of participants.

| Characteristic | AIP Risk Level |  |  |  | P value |
| --- | --- | --- | --- | --- | --- |
|  | Extremely Low Risk<br>(AIP < -0.3) | Low Risk<br>(-0.3 ≤ AIP < 0.1) | Medium Risk<br>(0.1 ≤ AIP < 0.24) | High Risk<br>(AIP ≥ 0.24) |  |
| Age, years,<br>Mean ± SE | 45.85 ± 17.24 | 47.85 ± 17.40 | 49.27 ± 16.67 | 49.13 ± 14.79 | <0.0001 |
| Gender, n (%) |  |  |  |  | <0.0001 |
| Male | 721 (36.04) | 479 (46.42) | 1850 (50.15) | 969 (64.82) |  |
| Female | 1279 (63.96) | 552 (53.58) | 1839 (49.85) | 526 (35.18) |  |
| Race, n (%) |  |  |  |  | <0.0001 |
| Mexican American | 114 (5.68) | 82 (7.94) | 429 (11.64) | 155 (10.37) |  |
| Non-Hispanic White | 1276 (63.78) | 724 (70.25) | 2512 (68.09) | 1085 (72.58) |  |
| Non-Hispanic Black | 323 (16.14) | 102 (9.94) | 192 (5.20) | 66 (4.43) |  |
| Others | 288 (14.4) | 122 (11.86) | 556 (15.07) | 189 (12.62) |  |
| Education level, n (%) |  |  |  |  | <0.0001 |
| High school or less | 595 (29.76) | 395 (38.29) | 1609 (43.62) | 645 (43.12) |  |
| Some college | 598 (29.90) | 334 (32.35) | 1200 (32.53) | 457 (30.60) |  |
| College graduate or above | 807 (40.35) | 303 (29.36) | 880 (23.85) | 393 (26.28) |  |
| Marital status, n (%) |  |  |  |  | <0.0001 |
| Divorced/Separated/<br>Widowed | 355 (17.73) | 195 (18.93) | 746 (20.23) | 293 (19.63) |  |
| Married/Living with a partner | 1228 (61.40) | 642 (62.30) | 2406 (65.21) | 998 (66.73) |  |
| Never married | 417 (20.86) | 194 (18.77) | 537 (14.56) | 204 (13.64) |  |
| Poverty-to-income ratio, n (%) |  |  |  |  | <0.0001 |
| < 1.3 | 441 (22.06) | 245 (23.74) | 1083 (29.37) | 409 (27.35) |  |
| 1.3–3.5 | 773 (38.66) | 432 (41.89) | 1452 (39.35) | 582 (38.90) |  |
| > 3.5 | 786 (39.28) | 354 (34.38) | 1154 (31.28) | 505 (33.75) |  |
| Depression, n (%) |  |  |  |  | <0.0001 |
| No | 1877 (93.84) | 953 (92.45) | 3242 (87.88) | 1352 (90.46) |  |
| Yes | 123 (6.16) | 78 (7.55) | 447 (12.12) | 143 (9.54) |  |
| 12-month average daily alcohol consumption, Mean ± SE | 0.31 ± 0.63 | 0.24 ± 0.73 | 0.21 ± 0.75 | 0.18 ± 0.39 | <0.0001 |
| LE8 score, Mean ± SE | 77.29 ± 13.19 | 68.39 ± 13.33 | 61.46 ± 13.64 | 59.59 ± 13.50 | <0.0001 |
| CVH score, n (%) |  |  |  |  | <0.0001 |
| Low (0–49) | 54 (2.72) | 93 (9.02) | 755 (20.46) | 359 (24.02) |  |
| Moderate (50–79) | 997 (49.83) | 723 (70.13) | 2646 (71.73) | 1031 (68.99) |  |
| High (80–100) | 949 (47.45) | 215 (20.85) | 288 (7.81) | 105 (6.99) |  |
| Health behaviors score, Mean ± SE | 72.06 ± 18.65 | 66.05 ± 19.42 | 62.10 ± 20.21 | 62.92 ± 19.54 | <0.0001 |
| HEI-2015 diet score, Mean ± SE | 53.70 ± 12.30 | 50.41 ± 11.97 | 49.20 ± 11.66 | 49.23 ± 11.28 | <0.0001 |
| Diet score, Mean ± SE | 48.12 ± 32.47 | 39.33 ± 31.69 | 36.40 ± 31.49 | 36.35 ± 30.76 | <0.0001 |
| Physical activity score, Mean ± SE | 76.64 ± 39.19 | 69.17 ± 42.99 | 63.34 ± 45.24 | 66.73 ± 44.42 | <0.0001 |
| Nicotine exposure score, Mean ± SE | 78.20 ± 35.13 | 72.66 ± 38.33 | 66.85 ± 39.92 | 65.32 ± 40.56 | <0.0001 |
| Sleep health score, Mean ± SE | 85.27 ± 22.66 | 83.06 ± 24.94 | 81.83 ± 25.24 | 83.29 ± 23.82 | 0.0008 |
| Health factors score, Mean ± SE | 82.52 ± 15.86 | 70.72 ± 17.71 | 60.83 ± 17.89 | 56.25 ± 17.76 | <0.0001 |
| Body mass index score, Mean ± SE | 76.99 ± 30.25 | 58.77 ± 33.54 | 46.19 ± 33.24 | 47.21 ± 30.69 | <0.0001 |
| Blood lipids score, Mean ± SE | 84.46 ± 22.12 | 68.76 ± 28.16 | 54.94 ± 29.09 | 41.45 ± 28.87 | <0.0001 |
| Blood glucose score, Mean ± SE | 90.64 ± 20.24 | 84.48 ± 24.07 | 77.16 ± 28.66 | 75.14 ± 30.49 | <0.0001 |
| Blood pressure score, Mean ± SE | 77.98 ± 29.77 | 70.88 ± 32.54 | 65.01 ± 32.70 | 61.21 ± 32.98 | <0.0001 |

AIP, Atherogenic index of Plasma; LE8, Life's Essential 8; HEI, healthy eating index; CVH:
Cardiovascular Health.

### 3.2 Distribution Relationship Between CVH Scores and AIP Risk Levels

**Figure 1** presented as a stacked bar chart, illustrates the percentage distribution of AIP risk levels among individuals with different CVH scores. The result showed within the low CVH score group, individuals at high AIP risk level comprised the largest proportion. As CVH scores increased, the proportions of individuals at high and medium AIP risk levels steadily decreased, while those at extremely low and low risk levels significantly rose. Most individuals with moderate CVH scores were classified within the low AIP risk level, whereas over 90% of individuals with high CVH scores fell into the extremely low or low risk categories.

**Figure 1.**
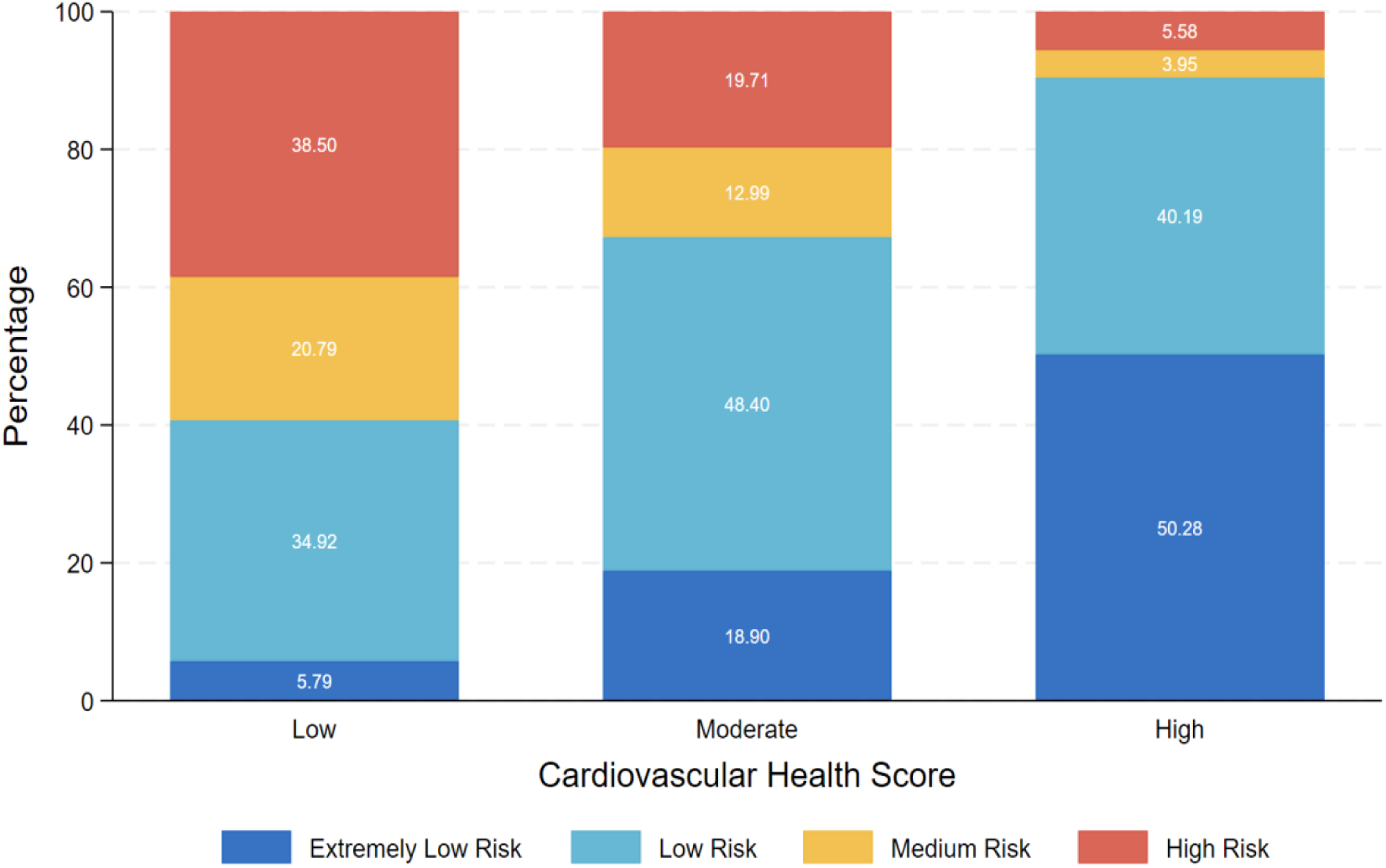
Stacked Bar Chart of CVH Scores and AIP Risk Levels.

### 3.3 Association between CVH Scores and AIP Risk Levels

The weighted ordered logistic regression analysis results summarized in **Table 2** indicated that the CVH score is significantly negatively associated with the AIP risk level. In the univariable analysis, every 10-point increase in the CVH score was associated with an approximately 44% decrease in the probability of an individual moving up one AIP risk level (*OR*=0.56, *95%CI*: 0.53-0.58, *P*<0.001). Compared to the low CVH score group, the probabilities of advancing one AIP risk level for the moderate and high CVH score groups decreased by approximately 65% (*OR*=0.35, *95%CI*: 0.29-0.41, *P*<0.001) and 92% (*OR*=0.08, *95%CI*: 0.06-0.10, *P*<0.001), respectively. This finding was further confirmed in the multivariable analysis, which showed that even after adjusting for age, sex, race, poverty-to-income ratio, education level, marital status, depression, and average daily alcohol consumption, the CVH score remained an independent predictor of the AIP risk level. In the multivariable analysis, for every 10-point increase in the CVH score, the probability of an individual moving up one AIP risk level decreased by approximately 49% (*OR*=0.51, *95%CI*: 0.49-0.54, *P*<0.001). Compared to the low CVH score group, the probabilities of advancing one AIP risk level decreased by approximately 69% for the moderate CVH score group (*OR*=0.31, *95%CI*: 0.26-0.37, *P*<0.001) and by 93% for the high CVH score group (*OR*=0.07, *95%CI*: 0.05-0.09, *P*<0.001). Additionally, both the health behavior score and the health factors score exhibited a significant negative correlation with the AIP risk level. In the univariable ordinal logistic regression analysis, each 10-point increase in the health behaviors score was associated with a 15% decrease in the probability of an individual advancing one AIP risk level (*OR*=0.85, *95%CI*: 0.83-0.88, *P*<0.001). For health factors scores, this probability decreased by approximately 39% (*OR*=0.61, *95%CI*: 0.59-0.63, *P*<0.001). In the multivariable ordinal logistic regression analysis, each 10-point increase in the health behaviors score corresponded to a 14% decrease in the probability of advancing one AIP risk level (*OR*=0.86, *95%CI*: 0.83-0.89, *P*<0.001). For health factors scores, the probability decreased by approximately 45% (*OR*=0.55, *95%CI*: 0.53-0.57, *P*<0.001). **Figure 2** illustrated the adjusted ORs and 95%CIs for the association between the CVH score and the AIP risk level across different subgroups. The result indicated that, in all subgroups, an increase in the CVH score was significantly associated with a reduction in the AIP risk level. This trend was validated across various subgroups defined by sex, race, education level, marital status, poverty-to-income ratio, and depression symptoms. Notably, the reduction in AIP risk level associated with a higher CVH score was particularly pronounced among females, non-Hispanic Black individuals, and individuals with depression symptoms.

**Table 2.** Association between CVH Score and AIP.

| Variables | Univariable model |  | Multivariable model |  |
| --- | --- | --- | --- | --- |
|  | OR (95% CI) | P value | OR (95% CI) | P value |
| CVH score |  |  |  |  |
| Per 10-point increase | 0.56 (0.53, 0.58) | < 0.001 | 0.51 (0.49, 0.54) | < 0.001 |
| Low (0–49) | Reference |  | Reference |  |
| Moderate (50–79) | 0.35 (0.29, 0.41) | < 0.001 | 0.31 (0.26, 0.37) | < 0.001 |
| High (80–100) | 0.08 (0.06, 0.10) | < 0.001 | 0.07 (0.05, 0.09) | < 0.001 |
| Health behaviors score |  |  |  |  |
| Per 10-point increase | 0.85 (0.83, 0.88) | < 0.001 | 0.86 (0.83, 0.89) | < 0.001 |
| Low (0–49) | Reference |  | Reference |  |
| Moderate (50–79) | 0.73 (0.63, 0.84) | < 0.001 | 0.75 (0.64, 0.89) | < 0.001 |
| High (80–100) | 0.46 (0.38, 0.55) | < 0.001 | 0.50 (0.41, 0.60) | < 0.001 |
| Health factors score |  |  |  |  |
| Per 10-point increase | 0.61 (0.59, 0.63) | < 0.001 | 0.55 (0.53, 0.57) | < 0.001 |
| Low (0–49) | Reference |  | Reference |  |
| Moderate (50–79) | 0.36 (0.31, 0.43) | < 0.001 | 0.32 (0.27, 0.38) | < 0.001 |
| High (80–100) | 0.08 (0.07, 0.10) | < 0.001 | 0.06 (0.05, 0.08) | < 0.001 |

OR, odds ratio; CI, confidence interval; CVH, cardiovascular health; LE8, Life's Essential 8.
Univariable model: unadjusted model.
Multivariable model: adjusted for age, sex, race, poverty-to-income ratio, education levels, marital status, depression, and 12-month average daily alcohol consumption.

**Figure 2.**
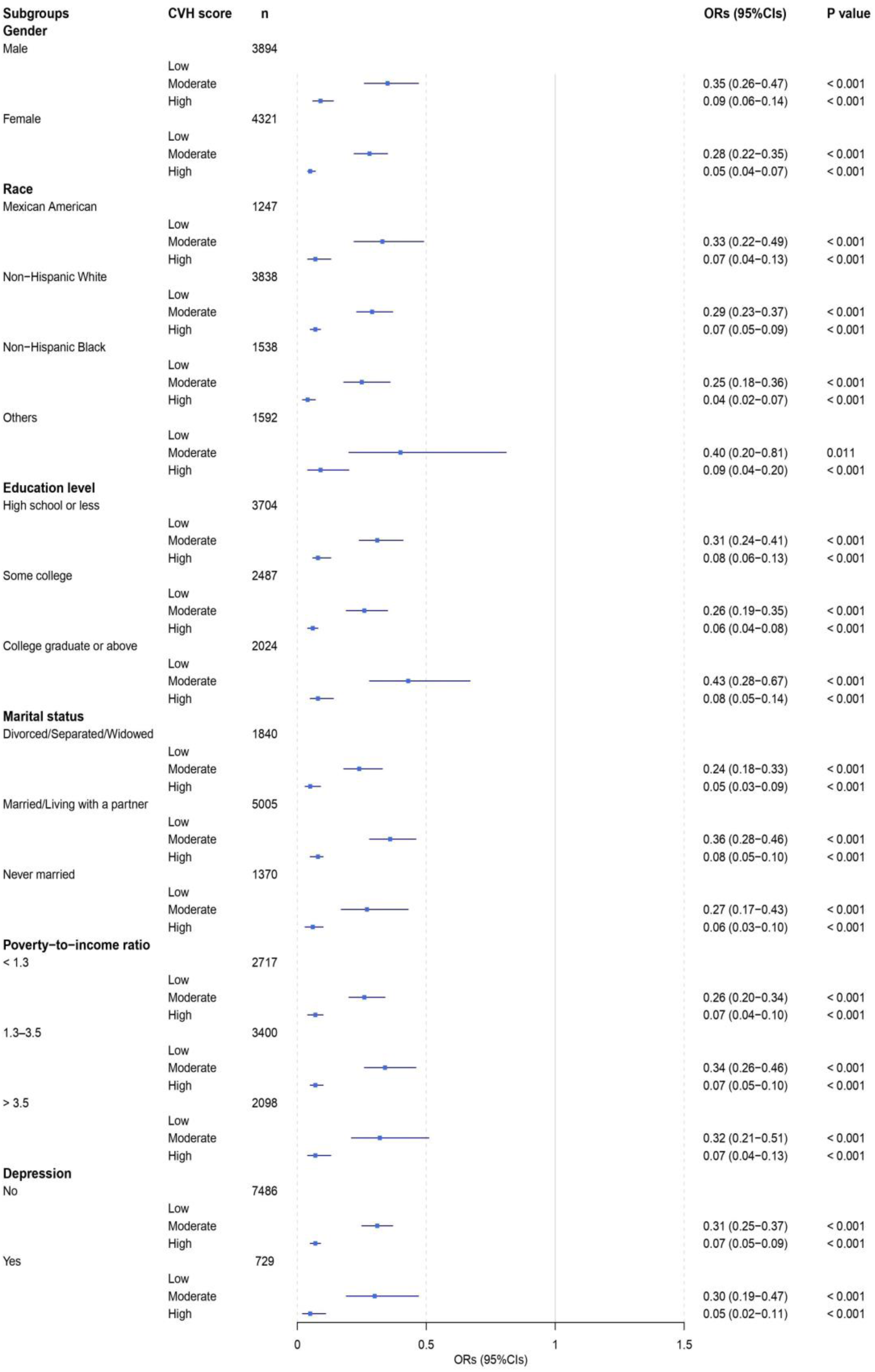
Adjusted ORs (95% CI) of AIP by subgroup of CVH metrics. ORs were adjusted for all listed covariates except the stratification variable in each subgroup analysis.

### 3.4 Association between individual CVH components and AIP

The result presented in **Figure 3** illustrate the adjusted ORs and 95%CIs for the association between different levels (low, moderate, high) of individual CVH components and the AIP risk level. The finding indicated that, with the exception of sleep health, improvements in all other CVH components were significantly associated with a reduction in AIP risk level, with the most notable effects observed for improvements in BMI and Blood Lipids scores. Notably, a high physical activity score was significantly associated with a lower AIP risk (*OR*=0.70, *95%CI*: 0.61-0.80, *P*<0.001). In contrast, the result for moderate physical activity scores did not achieve statistical significance (*OR*=0.79, *95%CI*: 0.59-1.06, *P*=0.117).

**Figure 3.**
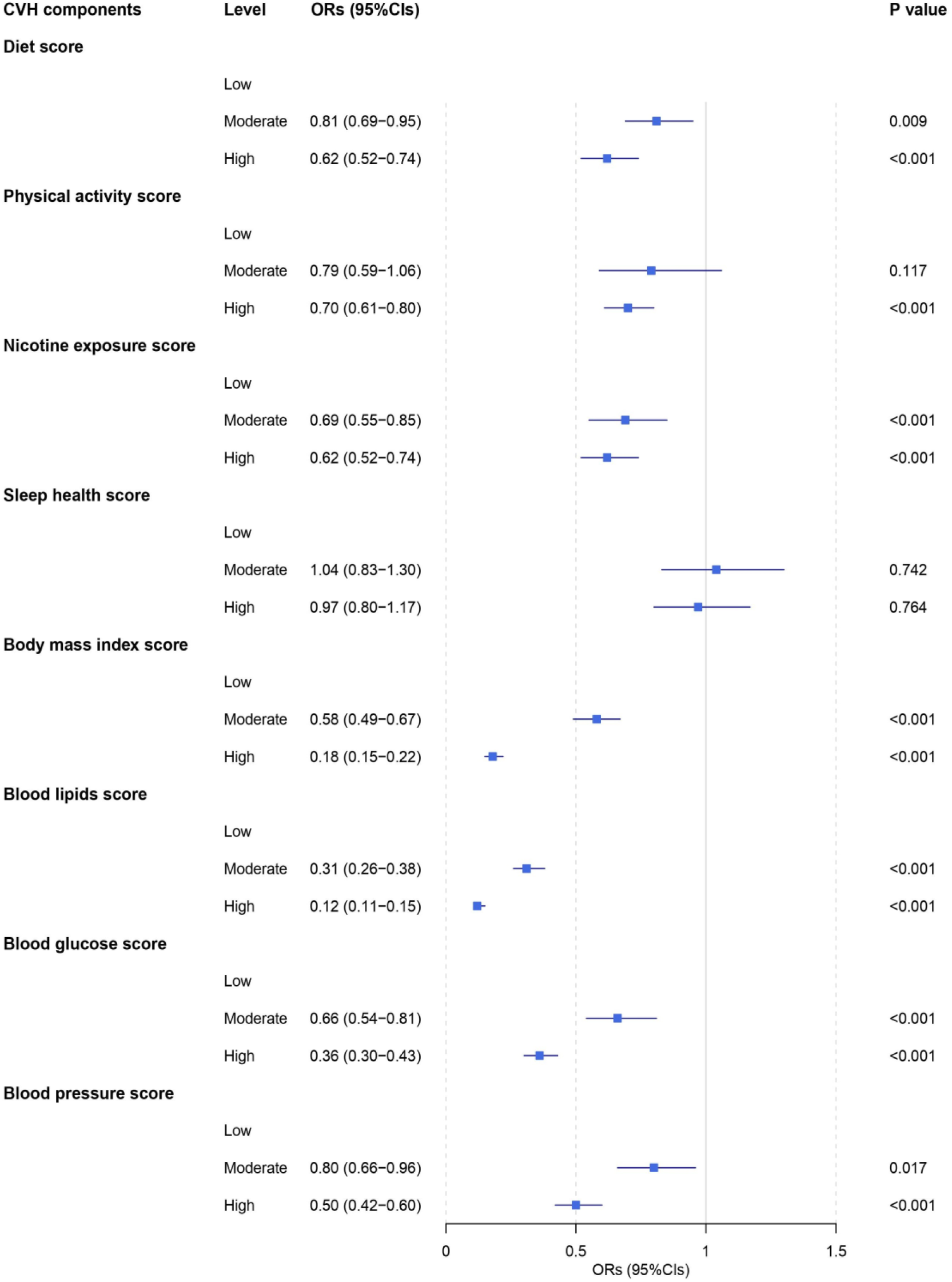
The association of CVH components with the AIP. ORs were adjusted for age, sex, race, poverty-to-income ratio, education levels, marital status, depression, and 12-month average daily alcohol consumption.

## 4. Discussion

To our knowledge, this study was the first to comprehensively explore the relationship between CVH and the AIP within the framework of LE8 using data from the NHANES database. The findings indicated a significant negative correlation between CVH scores and AIP risk levels: higher CVH scores were associated with lower AIP risk levels. Individuals with high CVH scores predominantly fell within the extremely low and low AIP risk levels, whereas those with low CVH scores were more frequently situated in the high AIP risk level. Furthermore, improvements in specific CVH components were correlated with reductions in AIP risk levels, particularly notable were the effects of enhancements in BMI and blood lipid levels.

The result of this study were consistent with those of Cunha et al.^24^, both demonstrating a negative correlation between CVH (LE8) scores and the AIP. Given that AIP is considered a sensitive predictor of CVD risk, it has exhibited excellent performance in identifying CVD risk factors: the area under the Receiver Operating Characteristic (ROC) curve was 0.909 (*P*<0.001), with an optimal cut-off value of 0.468, and high sensitivity (84.80%) and specificity (78.60%).^25^ The results of the this study may suggest that higher CVH scores are associated with lower CVD risk. A meta-analysis by Sebastian et al.^26^ further validated this phenomenon, revealing that the CVH score within the LE8 framework was a protective factor against CVD risk (*HR=*0.47, *95%CI*: 0.39-0.56, *P*<0.001), and individuals with high CVH scores had a lower all-cause mortality rate (*HR*=0.54, *95%CI*: 0.43-0.69, *P*<0.001) and disease-specific mortality rate (*HR*=0.37, *95%CI*: 0.26-0.52, *P*<0.001). Furthermore, despite the differences between the LE8 and LS7 frameworks, the CVH score remained significantly negatively correlated with AIP within the LS7 framework.^27,28^ This may indicate that the negative correlation between CVH and AIP possesses strong stability and universality, further enhancing the reliability of CVH scores as indicators for the prevention and management of CVD risk.

This study revealed that, with the exception of sleep health indicators, the remaining seven CVH metrics—namely Diet, Physical Activity, Nicotine Exposure, BMI, Blood Lipids, Blood Glucose, and Blood Pressure—exhibited significant correlations with the AIP risk levels. Notably, the improvements in BMI and blood lipid levels were particularly pronounced. Research conducted by Chang et al.^28^ similarly confirmed the associations of these seven metrics with AIP and indicated that BMI exerted the most substantial influence on AIP (*PR*=3.76; *95%CI*: 3.27-4.32; *P*<0.001). Analogously, the logistic regression analysis conducted by Shen et al.^29^ revealed that smoking status, BMI, physical activity, sodium intake, total cholesterol, blood pressure, and fasting blood glucose (FBG) all exhibited significant impacts on AIP (*P*<0.05), with BMI demonstrating the strongest correlation with total cholesterol and AIP. Furthermore, based on patterns of BMI change, Cao et al.^30^ discovered that an increase of over 10 kg in weight during the age span of 20 to 40 years was associated with a 22% increase in the risk of cardiovascular events (*HR*=1.22; *95%CI*: 1.04-1.43) and a 38% increase in the risk of diabetes (*HR*=1.38; *95%CI*: 1.25-1.53), when compared to stable weight. This may be attributed to the fact that elevated BMI is typically associated with abnormal or excessive fat accumulation, particularly in the visceral adipose tissue. Visceral fat is closely linked to metabolic syndrome (as increased visceral fat releases free fatty acids into the portal vein, becoming the primary source of hepatic TG synthesis, leading to elevated TG levels), insulin resistance (the rise in free fatty acids can suppress glucose transport or phosphorylation in muscle, thereby inducing insulin resistance), and inflammatory responses (the increased secretion of factors such as tumor necrosis factor-α and interleukin-6 leads to dysregulation of adipokine secretion), all of which contribute to elevated TG and reduced HDL-C, directly resulting in an increased AIP.^31–34^ Furthermore, previous research has revealed that obese adolescents exhibit significantly higher AIP levels compared to healthy controls, and a positive correlation between AIP and BMI has also been observed in adolescents with non-alcoholic fatty liver disease.^35^ However, this study only included participants aged 20 years and above; future investigations would benefit from expanding the age range or focusing on the younger population to further elucidate the dynamic changes in the relationship between BMI and AIP across different age groups.

Within the LE8 framework, sleep health is a multidimensional construct encompassing duration, timing, regularity, efficiency, satisfaction, and impact on daytime alertness. This metric is closely associated with CVD, mental health, and social determinants, and is of paramount importance for overall health management.^7^ A meta-analysis conducted by Behnoush et al.^36^ on three composite lipid indices— AIP, Lipid Accumulation Product (LAP), and Visceral Adiposity Index (VAI)—in patients with obstructive sleep apnea (OSA) revealed that AIP was significantly elevated in OSA patients, with the highest AIP values observed in those with severe OSA. This may be attributed to the fact that OSA patients, due to respiratory pauses and hypoxic events, are unable to obtain sufficient deep sleep, leading to inadequate sleep duration, which in turn triggers insulin resistance, elevated TG levels, and reduced HDL-C, ultimately resulting in an increase in AIP. However, this study found no statistically significant relationship between sleep health and AIP risk levels. This discrepancy may be attributed to the fact that this study primarily relied on participants’ self-reported average nightly sleep duration to calculate the sleep health score, which may have failed to comprehensively capture the multidimensional nature of sleep health. Future research should employ more comprehensive sleep health assessment metrics and incorporate objective monitoring devices, such as polysomnography or wearable technology, to record multiple dimensions of sleep duration, quality, and efficiency. This approach would further validate the relationship between sleep health and AIP risk levels, providing robust intervention strategies for CVH management.

Interestingly, the finding of this study revealed that the moderate level physical activity score (based on self-reported minutes of moderate or vigorous physical activity (MVPA) per week) did not exhibit statistical significance when compared to the low level physical activity score (*P*=0.117), whereas the high level physical activity score demonstrated statistical significance compared to the low level score (*P*<0.001). Previous research has shown a significant inverse dose-response relationship between physical activity and AIP: when MVPA was in the Q1 range of 0 MET·minutes/month, AIP exhibited a slight increase; when MVPA was in the Q2 range of 444-491 MET·minutes/month, AIP remained relatively unchanged; when MVPA was in the Q3 range of 2,457-2,596 MET·minutes/month, AIP decreased significantly; and when MVPA was in the Q4 range of 12,711-14,835 MET·minutes/month, AIP exhibited the most substantial reduction.^17^ Furthermore, Edwards et al.^37^ conducted additional research, revealing that the negative correlation between physical activity and AIP was entirely mediated by central obesity: without considering central obesity, MVPA was significantly associated with reduced odds of high AIP (*OR*=0.58, *95%CI*: 0.41-0.82, *P*=0.004); however, when central obesity was included as a covariate in the model, the association between MVPA and AIP was no longer significant (*OR*=0.80, *95%CI*: 0.55-1.18, *P*=0.260). In summary, the result of this study suggested that moderate level of physical activity may not have reached the critical threshold necessary to improve the dose-response relationship between central obesity and AIP, which may explain the lack of significant difference compared to low levels of physical activity. Conversely, high level of physical activity appear to effectively improve central obesity, thereby significantly reducing the AIP risk level. Future research should focus on identifying the optimal types and durations of physical activity that can enhance central obesity and AIP, providing a solid foundation for the development of more targeted and effective health management strategies.

This study had several limitations. First, the cross-sectional design of the study precluded the determination of causal relationships, and therefore, it could not be concluded that the improvement in CVH scores directly led to the reduction in AIP risk levels. Second, while the study adjusted for multiple confounding factors, there may have been potential unmeasured confounding variables that could have influenced the interpretation of the results. Third, some of the data (such as smoking, alcohol consumption, and dietary intake) relied on self-reports by the participants, which could have introduced information bias and affected the accuracy of the data. Fourth, the conversion of continuous variables into ordinal variables in this study may have resulted in the loss of information and reduction of details. Additionally, the determination of classification criteria involved a certain degree of subjectivity, which could have led to the incorrect classification of some individuals near the classification boundaries. Finally, this study primarily focused on AIP as an indicator of CVD risk, and it may have overlooked the comprehensive application of other relevant indicators and assessment methods.

## 5. Conclusion

This study has elucidated the significant inverse relationship between Life’s Essential 8 (LE8) and the Atherogenic Index of Plasma (AIP). The findings underscore the critical importance of enhancing cardiovascular health (CVH) scores in reducing AIP risk, thereby highlighting the essential role of health management strategies in the prevention of cardiovascular diseases (CVD). Despite the inherent limitations of this study, including the inability to establish causal relationships, the results offer valuable insights for the management of CVH. Looking forward, subsequent investigations should focus on a more comprehensive exploration of the causal dynamics between CVH and AIP, while also striving to develop more effective intervention strategies. Such efforts are crucial for promoting overall individual health and significantly diminishing the risk of CVD.

## Data Availability

The data utilized in this study are derived from the National Health and Nutrition Examination Survey (NHANES): https://wwwn.cdc.gov/nchs/nhanes/Default.aspx, and the data used are publicly available. The corresponding author can be contacted for data.

## 6. Acknowledgments

We would like to thank the staff at the Center for Big Data Research in Health and Medicine, The First Affiliated Hospital of Shandong First Medical University & Shandong Provincial Qianfoshan Hospital, for their valuable contribution.

## 7. Sources of Funding

No external funding.

## 8. Declaration of Competing Interest

The Authors declare that there is no conflict of interest.

## Notes

### Competing Interest Statement

The authors have declared no competing interest.

### Author Declarations

All study protocols have been approved by the National Center for Health Statistics Research Ethics Review Board, and written informed consent was obtained from all participants.

